# COVID-19 Anxiety Among Frontline Nurses: Predictive Role of Organisational Support, Personal Resilience and Social Support

**DOI:** 10.1101/2020.07.16.20141069

**Authors:** Leodoro J. Labrague, Janet de los Santos

## Abstract

**Aim:** This study examines the relative influence of personal resilience, social support and organisational support in reducing COVID-19 anxiety in frontline nurses.

**Background:** Anxiety related to the COVID-19 pandemic is prevalent in the nursing workforce, potentially affecting nurses’ well-being and work performance. Identifying factors that could help maintain mental health and reduce coronavirus-related anxiety among frontline nurses is imperative. Currently, no studies have been conducted examining the influence of personal resilience, social support and organisational support in reducing COVID-19 anxiety among nurses.

**Methods:** This cross-sectional study involved 325 registered nurses from the Philippines using four standardised scales.

**Results:** Of the 325 nurses in the study, 123 (37.8%) were found to have dysfunctional levels of anxiety. Using multiple linear regression analyses, social support (*β* = −0.142, *p* = 0.011), personal resilience (*β* = −0.151, *p* = 0.008) and organisational support (*β* = −0.127, *p* = 0.023) predicted COVID-19 anxiety. Nurse characteristics were not associated with COVID-19 anxiety.

**Conclusions:** Resilient nurses and those who perceived higher organisational and social support were more likely to report lower anxiety related to COVID-19.

**Implication for Nursing Management:** COVID-19 anxiety may be addressed through organisational interventions, including increasing social support, assuring adequate organisational support, providing psychological and mental support services and providing resilience-promoting and stress management interventions.

## Introduction

The coronavirus disease 2019 (COVID-19) pandemic is a substantial health burden that has major implications for public health globally. COVID□19 is a pneumonia-like disease caused by a novel coronavirus that emerged in the Province of Wuhan in China in November 2019. Worldwide, confirmed cases of the disease had reached 14,348,858 as of July 23, 2020, while in the Western Pacific Region, the confirmed cases had climbed to 263,565. Moreover, at this time there were 603,691 confirmed deaths due to COVID-19, and confirmed cases had been reported in more than 200 countries. The USA, Brazil, India, and Russia remain the countries with the highest numbers of confirmed cases, accounting for 49% of all confirmed cases globally (World Health Organization, 2020). Of the confirmed cases worldwide, 6%, or 90,000, were in healthcare workers. In the Philippines, since January 2020, the number of confirmed cases of COVID-19 has climbed to 70,764, with 45,646 active cases and 1837 confirmed fatalities. The country’s health agency identified 2736 healthcare workers who contracted the disease, of whom 1006 were nurses (Department of Health, 2020).

In a short span of time, COVID-19 has proven to be a fatal disease that has caused serious damage to the health and economy of the Philippines. The emergence of COVID-19 exerted unprecedented pressure on the country’s healthcare system and presented various challenges to its nursing workforce, potentially affecting nurses’ work performance and mental health, and even putting their lives at risk (Maben & Bridges, 2020; Mo *et al*., 2020; Ly *et al*., 2020).

## Review of Literature

Disease outbreaks such as the COVID-19 pandemic are anxiety-provoking situations. Defined as a “state of uneasiness or apprehension resulting from the anticipation of a real or perceived threatening event or situation” (Spielberger, 2010), anxiety is common among healthcare workers who are directly involved in managing affected patients during pandemics. Further, due to their direct contact with COVID-19 patients, healthcare workers (HCWs) are more exposed to traumatic events such as patients’ suffering and deaths (Pappa *et al*., 2020), which could further amplify their fears and anxiety. Available data suggest that the prevalence of anxiety among HCWs during pandemic ranged from 22.6% to 36.3% (Liu *et al*., 2020), rates that were significantly higher than those observed in the general population. Among HCWs, nurses were reported to experience the highest anxiety levels and the highest prevalence of anxiety, ranging from 15% to 92% (Alwani *et al*., 2020; Luo *et al*., 2020).

The main source of anxiety in nurses during the COVID-19 pandemic was fear of becoming infected or unknowingly infecting others (Mo *et al*., 2020). Shanafelt *et al*. (2020) identified other sources of anxiety in nurses, including lack of personal protective equipment (PPE), fear of harbouring the novel coronavirus at work, lack of access to COVID-19 testing, fear of transmitting the virus at work, doubt that their institution would support them if they became infected, lack of access to childcare facilities during lockdown, fear of being deployed in an unfamiliar ward or unit and lack of accurate information on the disease.

While a low level of anxiety is helpful to motivate and generate excitement in an individual, persistent exposure to anxiety may have negative consequences on their physio-psychological health and work performance. A vast number of studies have highlighted the negative effects of a higher levels of anxiety, including loss of desire to eat, dizziness, sleep disturbance and vomiting or nausea (Lee, 2020; Lee *et al*., 2020). Higher anxiety levels were also associated with impairment in some bodily functions, negative coping mechanisms (such as increased intake of alcohol or drugs), stress and depression and increased suicidal ideation (Lee *et al*., 2020). Further, unmanaged anxiety may lead to long-term effects on nurses’ work performance and job satisfaction, leading to frequent absenteeism and eventual turnover (Labrague *et al*., 2018a; Lee *et al*., 2020). Implementing measures to reduce anxiety among nurses may prevent its adverse consequences. Such measures are vital to sustaining a well-engaged nursing workforce.

Nurse managers play a vital role in addressing nurses’ anxiety or fears of COVID-19 by supporting their mental, psychological and emotional health through evidence-based measures, supportive organisational policies and provision of a safe and secure work environment (Mo *et al*., 2020; Catton, 2020). Personal resilience and social and organisational support were identified in the literature as vital factors protecting against adversity and stress in nurses, allowing them to maintain their mental well-being and psychological health (Labrague *et al*., 2018b; Turner, 2015; Kim & Park, 2017; Bloom *et al*., 2017).

Personal resilience, or a person’s capacity to ‘bounce back’ or recover quickly from a stressful event (Hart *et al*., 2014), may help nurses cope effectively and endure the burden caused by stressors. In the context of the COVID-19 pandemic, personal resilience may help nurses effectively endure the stress caused by the pandemic (Cooper *et al*., 2020). Available studies identified the protective role of personal resilience in nurses during disaster events (Labrague *et al*., 2018b; Turner, 2015) and disease outbreak (Duncan, 2020), suggesting that strengthening nurses’ levels of hardiness and coping abilities can help them manage and deal with stressful situations effectively.

Defined as the assistance and protection given to others, especially individuals (Langford *et al*., 1997), social support drawn from colleagues, managers, friends and families is considered to be important for nurses to cope and deal effectively with different stressors in the work environment. A wide range of studies identified the positive effects of social support on nurses’ job satisfaction, work commitment, health and well-being (Choi, 2018; Hu *et al*., 2018).

Adequate social support was also seen as vital to helping healthcare workers effectively manage stressful events, including emergency situations, disaster events and outbreak of infectious diseases (Labrague *et al*., 2018c; Kim & Park, 2017).

Organisational support, or the degree to which an organisation provides resources, reinforcement, encouragement and communication to an individual to perform their functions effectively, is a vital factor that contributes to organisational success (Eisenberger *et al*., 1986, 2002). Mounting evidence has shown a positive link between higher levels of organisational support and positive outcomes in nurses (e.g. work performance, job satisfaction, innovative behaviours) and patients (e.g. patient satisfaction) (Labrague *et al*., 2018c; Pahlevan Sharif *et al*., 2018). Evidence has also shown that higher levels of organisational support may reduce the impact of the different workplace stressors and may serve as a protective factor against stress and anxiety caused by disasters, calamities and other emerging infectious diseases (Bloom *et al*., 2017; Veenema *et al*., 2017).

Despite the abundance of studies examining the importance of personal resilience, social support and organisational support in helping nurses maintain health and well-being during stressful events, no studies have yet been conducted to examine how these elements contribute to the reduction of anxiety related to COVID-19. Hence, this study was conducted to assess the causal relationships between personal resilience, social support, organisational support and COVID-19 anxiety.

## Methods

### Research Design

A cross-sectional research design was adopted in this study.

### Samples and Settings

This study was conducted in the Philippines, a Southeast Asian country with a population of 104.9 million, making it the 13th-most populous nation in the world (Philippine Statistics Authority, 2018). The country’s physical health infrastructure is comprised of 1224 hospitals (434 government hospitals and 790 private hospitals) distributed throughout the 17 Regions in the country (Dayrit *et al*., 2018). This study was conducted in one Region in the Philippines, Region 8, which is comprised of 50 government and 25 privately-owned hospitals. As of 2018, there are approximately 3000 nurses working all hospitals within the Region, with most nurses employed in government hospitals. As a consequence of increased migration of experienced nurses to other countries, such as the UK, the USA and countries in the Middle East, the current nurse workforce in the Region is composed mainly of young nurses with an average age of 27 to 35 years (Baustista *et al*., 2020; Labrague *et al*., 2020).

In this study, licensed registered nurses (RNs) assigned to 75 wards or units from 10 government and 10 private hospitals within the Region were recruited to take part in the study. These hospitals were randomly selected from a list of all hospitals in the Region. Inclusion criteria for the hospitals were: a minimum of 50 beds, an emergency department and an operating department and provision of health services to COVID-19 patients. To achieve an 80% power, with alpha set at 0.05 and small effect size set at 0.05 (Soper, 2020), a sample size of 293 nurses would be required, as determined using the G*power program, software version 3.1.9.9 (Faul *et al*., 2007). The sample size was initially calculated using power estimates for seven predictors in multiple regression. A total of 350 nurses were initially invited to partake in the study; however, only 325 nurses responded (93% response rate). A two-phase sampling approach was followed during the recruitment of the participants: stratified sampling in determining the number of nurses per hospital (Phase 1) and convenience sampling to select the respondents in each hospital (Phase 2). Participants were included if they were (a) a registered nurse, (b) held either a permanent or contracted/casual role, (c) were currently employed in either a private or a public hospital and (d) had more than six months of experience working as a nurse.

### Instrumentation

Four standardised, self-reported scales were used for data collection: the COVID-19 Anxiety Scale, the Brief Resilient Coping Scale (BRCS), the Perceived Social Support Questionnaire (PSSQ) and the Perceived Organizational Support (POS) questionnaire. The original English versions of the scales were used in this study, as nurses in the country are proficient in the English language.

To assess or identify individuals who may have abnormal levels of anxiety related to the COVID-19 pandemic, the COVID-19 Anxiety Scale, which was designed and developed by Lee *et al*. (2020), was used. This scale contains five items that reflect the common symptoms of anxiety experienced by an individual and was originally designed to examine anxiety over COVID-19 in 775 adults in the USA. Possible scores for this scale ranged from 5 to 25. The scale discriminates between those with dysfunctional anxiety and non-anxiety using an optimised cut-off score of 9 (Lee *et al*., 2020). Dysfunctional anxiety refers to a disproportionate state of anxiety, defined as persistent or uncontrollable fear that interferes with daily life and causes disruptions to behaviour and psychological well-being (Lee *et al*., 2020). Nurses participating in the study indicated the frequency of symptoms experienced in a 5-point Likert-type scale (0 [not at all] to 4 [nearly every day]). The scale had an outstanding predictive validity, as evidenced by a positive association with disability and psychological distress (Lee *et al*., 2020), and excellent reliability, with an internal consistency of 0.93 in a previous study (Lee *et al*., 2020) and a Cronbach’s alpha of 0.87 in the present study.

To examine nurses’ ability to rebound from stressful events, the BRCS, which was developed by Smith *et al*. (2008), was utilised. To complete the BRCS, nurses indicated their responses on each of the four items using a 5-point Likert-type scale (0 [does not describe me at all] to 5 [describes me very well]). The mean scale scores were categorised into three groups: low resilience (1.00–2.99), moderate resilience (3.00–4.30) and high resilience (4.31–5.00; Smith *et al*., 2008). The scale demonstrated an outstanding predictive validity, as evidenced by its positive association with work performance, health and well-being (Foster *et al*., 2019), and an excellent reliability, with an internal consistency of 0.91 in previous research (Smith *et al*., 2008) and a Cronbach’s alpha of 0.84 in the current study.

To assess nurses’ perceptions of the extent of support they receive from others when facing stressful situations, the PSSQ developed by Lin *et al*. (2019) was used. The PSSQ is a six-item scale that the nurses answered by indicating the extent of their agreement with each item using a 5-point Likert-type scale (1 [strongly disagree] to 5 [strongly agree]). The mean scale scores were divided into three categories: low social support (1.00–2.99), moderate social support (3.00–4.30) and high social support (4.31–5.00; Lin *et al*., 2019). The scale had excellent criterion validity, demonstrated by its negative correlation with emotional exhaustion and turnover intention (Duan *et al*., 2019), and excellent reliability, with an internal consistency of 0.89 in a previous study (Lin *et al*., 2019) and a Cronbach’s alpha of 0.86 in the present study.

To assess nurses’ opinions on the extent to which their workplace recognises and values their well□being, the POS scale, which was developed and designed by Eisenberger *et al*. (1986), was utilised. The POS is an eight-item scale on which nurses indicated the extent of their agreement with each item using a 5-point Likert□type scale (1 [strongly disagree] to 5 [strongly agree]). The mean scale scores were categorised as follows: low organisational support (1.00– 2.99), moderate organisational support (3.00–4.30) and high organisational support (4.31–5.00; Labrague *et al*., 2018). The scale had an excellent criterion validity, as evidenced by its positive association with job satisfaction, quality of life and psychological well-being (Labrague *et al*., 2018), and excellent reliability, with an internal consistency of 0.93 in a previous study (Eisenberger *et al*., 1986) and a Cronbach’s alpha of 0.87 in the present study.

### Ethical Considerations and Data Collection

Data collection using a questionnaire was conducted after ethical clearance was granted by the Institutional Research Ethics Committee of Samar State University. Letters asking for permission to collect data were sought from the nurse executives of the selected hospitals a few days prior to the conduct of the study. After the potential participants were identified by the researchers and/or trained researcher assistants based on the inclusion criteria, a short orientation was given, written consent was sought and the questionnaires were handed to the participants in a sealed envelope by a researcher or research assistant. Participants were asked to complete the questionnaire during their break time so as not to disrupt their work. Data were collected from April 25 to May 25, 2020.

### Data Analysis

Means, frequencies and standard deviations (SDs) were used to quantify the data. Nurse variables and key study variables were correlated to COVID-19 anxiety using Student’s t-test, Pearson’s correlation coefficient and analysis of variance. Variables that correlated significantly with the dependent variable were entered into the regression model (enter method) to identify potential predictors of COVID-19 anxiety. Data were considered statistically significant if the p-value for a particular statistical test was < 0.05. Data were analysed using version 23 of SPSS Statistics software for Windows 22.

## Results

A total of 325 nurses responded to the questionnaires. Their mean age was 30.94 years (SD = 6.67), while the average years in the current organisation and in the nursing profession were 4.65 years and 8.92 years, respectively. The majority of the respondents were female (74.8%), unmarried (66.8%) and held a baccalaureate nursing degree (82.2%). The detailed characteristics of the nurses are presented in **Table 1**. Regarding preparedness to care for COVID-19 patients, 28 (8.6%) reported that they were ‘prepared’, 104 (32%) were ‘somewhat prepared’, and nearly half (45.2%) were ‘unsure’. When asked if they were willing to care for COVID-19 patients, 26.8% answered ‘probably yes’, 20.3% answered ‘absolutely yes’ and a greater proportion were ‘unsure’ (35.7%) or ‘not willing’ (17.2%).

**Table 1.**
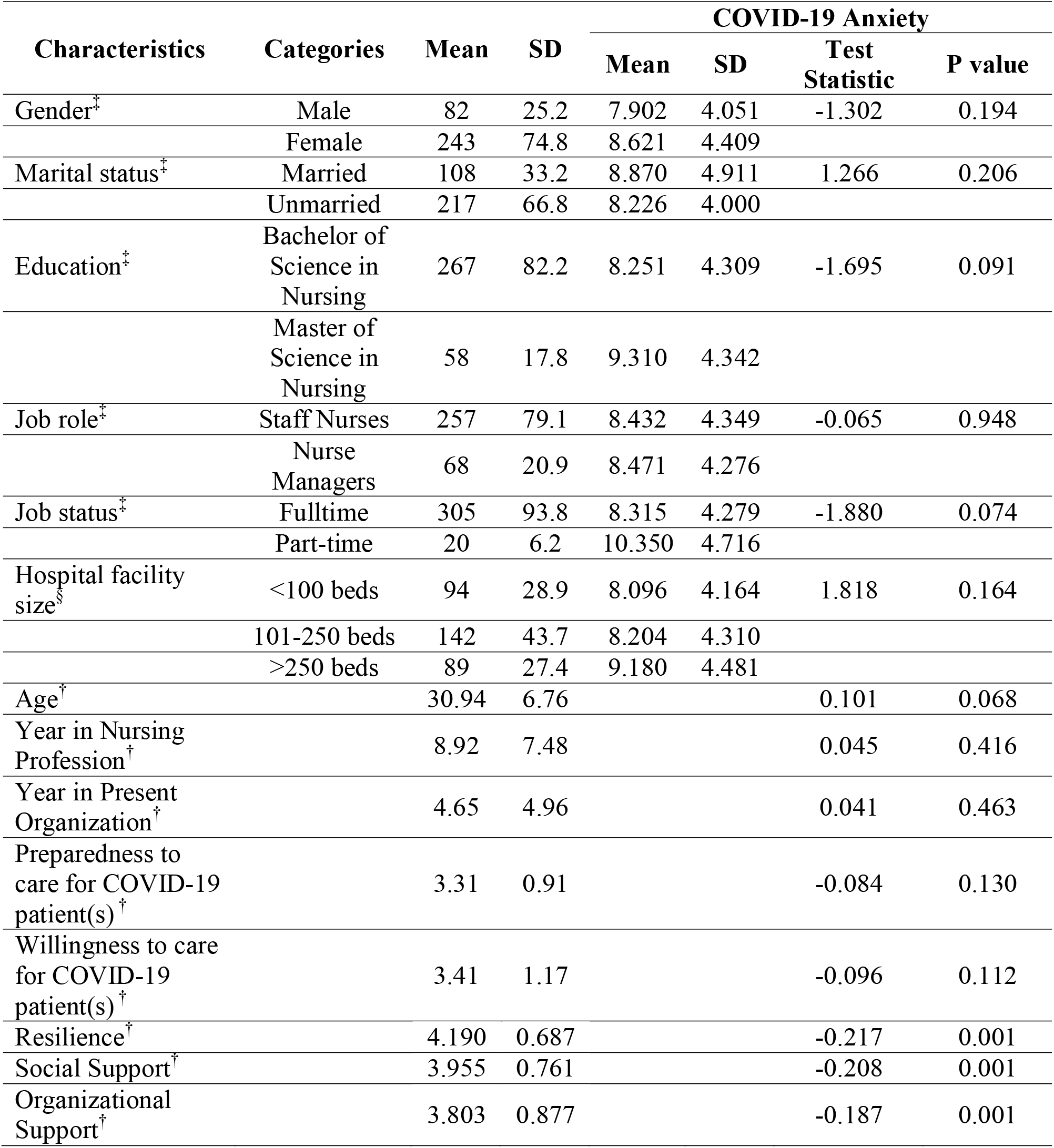
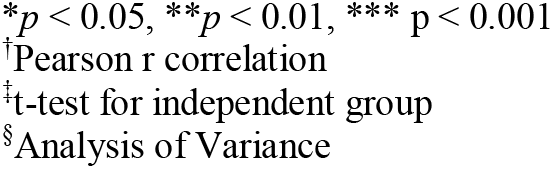
Nurses’ demographic and professional information and its relationship with COVID-19 anxiety mean score

| Characteristics | Categories | Mean | SD | COVID-19 Anxiety |  |  |  |
| --- | --- | --- | --- | --- | --- | --- | --- |
|  |  |  |  | Mean | SD | Test Statistic | P value |
| Gender<sup>‡</sup> | Male | 82 | 25.2 | 7.902 | 4.051 | -1.302 | 0.194 |
|  | Female | 243 | 74.8 | 8.621 | 4.409 |  |  |
| Marital status<sup>‡</sup> | Married | 108 | 33.2 | 8.870 | 4.911 | 1.266 | 0.206 |
|  | Unmarried | 217 | 66.8 | 8.226 | 4.000 |  |  |
| Education<sup>‡</sup> | Bachelor of Science in Nursing | 267 | 82.2 | 8.251 | 4.309 | -1.695 | 0.091 |
|  | Master of Science in Nursing | 58 | 17.8 | 9.310 | 4.342 |  |  |
| Job role<sup>‡</sup> | Staff Nurses | 257 | 79.1 | 8.432 | 4.349 | -0.065 | 0.948 |
|  | Nurse Managers | 68 | 20.9 | 8.471 | 4.276 |  |  |
| Job status<sup>‡</sup> | Fulltime | 305 | 93.8 | 8.315 | 4.279 | -1.880 | 0.074 |
|  | Part-time | 20 | 6.2 | 10.350 | 4.716 |  |  |
| Hospital facility size<sup>§</sup> | <100 beds | 94 | 28.9 | 8.096 | 4.164 | 1.818 | 0.164 |
|  | 101-250 beds | 142 | 43.7 | 8.204 | 4.310 |  |  |
|  | >250 beds | 89 | 27.4 | 9.180 | 4.481 |  |  |
| Age<sup>†</sup> |  | 30.94 | 6.76 |  |  | 0.101 | 0.068 |
| Year in Nursing Profession<sup>†</sup> |  | 8.92 | 7.48 |  |  | 0.045 | 0.416 |
| Year in Present Organization<sup>†</sup> |  | 4.65 | 4.96 |  |  | 0.041 | 0.463 |
| Preparedness to care for COVID-19 patient(s)<sup>†</sup> |  | 3.31 | 0.91 |  |  | -0.084 | 0.130 |
| Willingness to care for COVID-19 patient(s)<sup>†</sup> |  | 3.41 | 1.17 |  |  | -0.096 | 0.112 |
| Resilience<sup>†</sup> |  | 4.190 | 0.687 |  |  | -0.217 | 0.001 |
| Social Support<sup>†</sup> |  | 3.955 | 0.761 |  |  | -0.208 | 0.001 |
| Organizational Support<sup>†</sup> |  | 3.803 | 0.877 |  |  | -0.187 | 0.001 |

\* $p < 0.05$ , \*\* $p < 0.01$ , \*\*\* $p < 0.001$
<sup>†</sup>Pearson r correlation
<sup>‡</sup>t-test for independent group
<sup>§</sup>Analysis of Variance

Bivariate analysis showed no significant relationship between nurses’ variables and the composite score of the COVID-19 Anxiety Scale. Pearson’s correlation analysis revealed weak but significant negative relationships between COVID-19 anxiety and personal resilience (*r* = - 0.217, *p* > 0.001), social support (*r* = −0.208, *p* > 0.001) and organisational support (*r* = −0.187, *p* > 0.001; **Table 1**).

Regarding the descriptive statistics of other key study variables, the mean scale score on the Brief Resilient Coping Scale was 4.190 (SD = 0.687), which was interpreted as ‘normal resilience’. The mean scale scores for the Perceived Social Support Questionnaire and Perceived Organizational Support Scale were 3.955 and 3.803, which were interpreted as ‘moderate social support’ and ‘moderate organisational support’, respectively (**Table 1**). Based on the cut-off score of ≥ 9.0 in the COVID-19 Anxiety Scale (Lee *et al*., 2020), 123 respondents (37.8%) were found to have dysfunctional levels of anxiety. The mean scale score was 8.440 (SD = 4.32), with ‘tonic immobility’ (1.779) as the highest-rated item, followed by ‘sleep disturbance’ (1.726) and ‘dizziness’ (1.655; **Table 2**).

**Table 2.** Descriptive statistics of the COVID-19 Anxiety Scale

| Scale/Subscale | n | Min | Max | Mean | SD |
| --- | --- | --- | --- | --- | --- |
| COVID-19 Anxiety<sup>†</sup> | 325 | 5.00 | 25.00 | 8.440 | 4.326 |
| Dizziness<sup>‡</sup> | 325 | 1.00 | 5.00 | 1.655 | 0.919 |
| Sleep Disturbance<sup>‡</sup> | 325 | 1.00 | 5.00 | 1.726 | 1.028 |
| Tonic Immobility<sup>‡</sup> | 325 | 1.00 | 5.00 | 1.779 | 1.027 |
| Appetite Loss<sup>‡</sup> | 325 | 1.00 | 5.00 | 1.643 | 0.989 |
| Abdominal Distress<sup>‡</sup> | 325 | 1.00 | 5.00 | 1.637 | 0.974 |

<sup>†</sup>mean scale score
<sup>‡</sup>mean item score

Based on multiple linear regression analyses, social support (*β* = −0.142, *p* = 0.011), personal resilience (*β* = −0.151, *p* = 0.008) and organisational support (*β* = −0.127, *p* = 0.023) predicted COVID-19 anxiety. In other words, increased scores in the social support, organisational support and personal resilience measures were associated with decreased scores in the COVID-19 Anxiety Scale scores (**Table 3**).

**Table 3.** Influence of social support, resilience, and organizational support on COVID-19 Anxiety

| Predictors | B | SE | $\beta$ | t | *p* value | CI |
| --- | --- | --- | --- | --- | --- | --- |
| Constant | 16.577 | 1.598 |  | 10.375 | 0.001 | 13.433 to 19.720 |
| Social Support | -0.523 | 0.203 | -0.142 | -2.572 | 0.011 | -0.923 to -0.123 |
| Resilience | -0.947 | 0.356 | -0.151 | -2.662 | 0.008 | -1.647 to -0.247 |
| Organizational Support | -0.628 | 0.274 | -0.127 | -2.291 | 0.023 | -1.116 to -0.089 |

**Note:**
Controlling for nurse/unit/hospital characteristics (age, year in the present unit, year in the organization, marital status, education, job role, facility size, hospital type)
$\beta$ , Standardized Regression Coefficient; SE, Standard Error; CI, Confidence Interval

## Discussion

Frontline nurses in the Philippines reported moderate levels of personal resilience and perceived moderate levels of social and organisational support during the COVID-19 pandemic. Moreover, increased levels of personal resilience, organisational support and social support in nurses were associated with decreased levels of anxiety related to COVID-19.

The resilience of frontline nurses in the current study was moderate. This finding is in accordance with international studies that identified nurses working in hospitals as having moderate levels of personal resilience (Guo *et al*., 2017; Kutluturkan *et al*., 2016). As personal resilience impacts work performance, health and overall well-being in nurses, it is vital to enhance this personal resource through proactive organisational measures. Further, frontline nurses in our study perceived moderate levels of social support and organisational support, which is consistent with earlier studies. Since higher levels of social and organisational support were significantly associated with positive work outcomes (e.g. work performance, job satisfaction, job engagement) and physical and mental health in nurses (Labrague *et al*., 2018c; Hu *et al*., 2018), it is critical that measures aimed towards improving these elements are implemented in the workplace.

More than 90% of frontline nurses reported that they were not fully prepared to manage COVID-19 patients, and only 20.3% reported being absolutely willing to care for COVID-19 patients. The proportion of nurses who expressed their willingness to manage patients affected by the COVID-19 outbreak was lower than in previous studies focused on other infectious diseases in which more than 75% of nurses expressed their willingness to care for patients affected by diseases such as H1NI and Ebola (McMullan *et al*., 2016; Bugade *et al*., 2018). In addition, this study corroborates earlier studies in which many nurses reported not being sufficiently prepared to handle patients affected by infectious disease (e.g. Ebola, H1N1; MnMullan *et al*., 2016; Bugade *et al*., 2018) or respond to emergency and disaster situations (Labrague *et al*., 2018b; Labrague *et al*., 2016; Tzeng *et al*., 2016). The results of this study highlight the need for nurses to be fully equipped with the required competencies in order to better handle and manage patients affected by disease outbreaks and in emergency situations.

Using a cut-off score of ≥ 9.0 on the COVID-19 Anxiety Scale, 37.8% of the respondents were identified to have dysfunctional levels of anxiety. Due to the apparent lack of studies using similar tools in which nurses were the participants, comparison with highly similar studies was not possible. However, the proportion of participants in this study who experienced dysfunctional levels of COVID-19 anxiety was lower than in the general population (54.8%; Lee *et al*., 2020). This may be due to the fact that nurses have wider knowledge of the nature of COVID-19, its transmission and symptoms and measures to prevent the disease than the general population. Since nurses are directly involved in the care of COVID-19 patients and the delivery of healthcare services, it is essential to implement measures to reduce anxiety levels among nurses, as dysfunctional anxiety levels have been identified as strong precursors of psychological distress, depression and other psychological disorders (Teles, Barbosa, & Vargas, 2014; Mo *et al*., 2020).

In this study, among the different symptoms of coronavirus anxiety, ‘tonic immobility’ and ‘sleep disturbance’ were reported to be the most pronounced symptoms. This result is similar to those of a study involving the general population in Poland in which these items obtained the highest means (Skalski *et al*., 2020). In addition, in a study by Shelvin *et al*. (2020), high levels of anxiety were linked to somatic symptoms such as fatigue and gastrointestinal manifestations. Accordingly, ‘tonic immobility’ and ‘sleep disturbance’ were identified in the literature as the most common responses to threatening events or situations. Sleep disturbance is a symptom common among individuals who have post-traumatic stress disorder and anxiety disorder (APA, 2013), while tonic immobility occurs involuntarily as a result of heightened fear and typically occurs in individuals who experience highly traumatic situations (Moller *et al*., 2017).

The most important findings of this study were the significant effects of personal resilience, social support and organisational support on COVID-19 anxiety levels in nurses, above and beyond the influence of nurse characteristics. The reduced COVID-19 anxiety levels in nurses who had higher scores on the resilience scale demonstrate the protective role of personal resilience, which enables an individual to positively adapt in stressful and anxiety-provoking situations and bounce back successfully despite adverse circumstances (Foster *et al*., 2019). This result highlights the importance of developing measures or interventions to promote or optimise personal resilience in frontline nurses in order to reduce their anxiety related to COVID-19. This result is in line with previous studies linking higher resilience in nurses to reduced burnout, compassion fatigue, anxiety, depression and psychological distress (Mealer *et al*., 2017; Cooper *et al*., 2020). Higher resilience was also associated with enhanced outcomes in an individual, such as increased psychological health and mental well-being (Foster *et al*., 2020; Gao *et al*., 2017). In a literature review by De Brier *et al*. (2020), maximising resilience in HCWs during the COVID-19 crisis was seen as vital to helping them safeguard their mental and psychological health and well-being. This pattern of influence was similar to that found in a general population, in which personal resilience and social support contributed significantly to reducing the severity of anxiety associated with the novel coronavirus (Skalski *et al*., 2020).

Available literature has identified social support that originates from colleagues, friends and families as effective support systems in nurses. Such support systems are essential when facing anxiety-provoking events. In this study, increased scores on the Perceived Social Support Questionnaire were associated with significantly lower COVID-19 Anxiety Scale scores. This result corroborates previous studies underlining the vital role of adequate social support in helping nurses achieve positive emotional states during stressful events such as disease outbreaks (Li & Aungsuroch, 2019). In a study involving doctors and nurses, higher scores on the social support scale were negatively associated with anxiety, depression and sleep disorders, suggesting that psychological symptoms of healthcare staff could be eased by enhancing social support during the COVID-19 pandemic (Zou *et al*., 2020). Further, positive coping strategies and increased social support were associated with decreased psychological distress, increased self-efficacy, improved sleep quality and decreased levels of anxiety and stress among nurses (Yu *et al*., 2020; Xiao *et al*., 2020).

Finally, increased scores on the organisational support measure were associated with decreased scores on the COVID-19 Anxiety Scale. This result was expected, as adequate organisational support, or the degree to which the organisation recognises employees and values their well-being, has been associated with increased job performance and commitment in nurses, both of which are vital when dealing with a disease outbreak (Shiao *et al*., 2007; Jung *et al*., 2020). Further, when nurses perceived greater support from the organisation, they were more motivated, were highly satisfied and experienced less stress when carrying out their duties (Higazee *et al*., 2016; Labrague *et al*., 2019c). This result suggests that during disease outbreaks such as the COVID-19 pandemic, when stress and anxiety are high, providing adequate organisational support (e.g. structural support, efficient communication, provision of a safe work environment, trainings related to COVID-19, monitoring of HCWs’ health and well-being) is vital to assist nurses facing the challenges brought about by the coronavirus crisis.

### Study Limitations

While this study provides evidence important for nursing administrators to assist nurses during pandemics, a few limitations were identified. Since this study involved nurses from only one province of the Philippines, results cannot be generalised to nurses throughout the country or the world. Next, the nature of the study design poses some limitations; for example, establishing causal links between variables may not be possible. The use of self-reported measures may have limited the responses of the participants; therefore, future studies may utilise both qualitative and quantitative designs to elicit essential information from the participants that could not have been captured by the scales used. Further, future studies utilising rigorous methods (e.g. experimental research design) may be conducted to examine and test the effectiveness of a resilience program and other initiatives to increase social and organisational support in reducing coronavirus anxiety in nurses. Finally, future research should explore how personal (e.g. self-efficacy, coping skills, hardiness) and organisational (adequacy of healthcare staffing, adequacy of hospital resources, number of patients admitted, hospital size) variables affect coronavirus anxiety in nurses.

### Implications for Nursing Management

Since dysfunctional levels of COVID-19 anxiety may have negative effects on nurses’ mental health and well-being, provision of adequate mental and psychological support should be prioritised by nurse managers, who should institute evidence-based measures to promote the mental health of nurses during the COVID-19 pandemic. Nurse managers should ensure that nurses are given access to psychological treatment or psychotherapy, as well as materials and resources to support mental health. Nurse managers should prioritise and promote self-care among nurses by offering flexible or shorter duty hours, adequate breaks and time scheduling, for example, which may help reduce the negative impact of the crisis and reduce nurses’ anxiety.

Nurse managers should also prioritise building personal resilience among frontline nurses, as higher personal resilience was associated with lower COVID-19 anxiety. By reinforcing positive coping strategies and supporting nurses’ self-efficacy, nurse managers can better promote resilience among nursing staff, which is essential when dealing with workplace adversity and stressful work situations such as disease outbreaks or pandemics. Social support, including support originating from colleagues, friends and families, may help provide a sense of security in nurses and help alleviate their fears during a pandemic. Through sharing of work experiences, listening to nurses’ concerns and offering empathetic support, nurses’ mental health and psychological well-being will be supported and their morale will improve. Nurse managers should provide adequate organisational support through the implementation of a safe work environment, provision of complete and quality PPE and supplies to prevent infection, provision of accurate and timely information regarding the disease and implementation of trainings relevant to COVID-19. These organisational practices are critically important to support nurses in their nursing practices and protect both their physical and mental health.

## Conclusion

The COVID-19 pandemic may cause dysfunctional levels of anxiety in frontline nurses. Increased levels of personal resilience, social support and organisational support were associated with decreased levels of anxiety related to the COVID-19 pandemic. Thus, organisational strategies to enhance personal resilience and increase social and organisational support in nurses may reduce their anxiety related to the COVID-19 pandemic, the management of which is critically important when caring for patients affected by COVID-19.

## Data Availability

Data are available upon request.

## Notes

### Competing Interest Statement

The authors have declared no competing interest.

### Funding Statement

This study is non-funded.

### Author Declarations

Public State University (PSU-2020-001-25)

